# Development of a severity of disease score and classification model by machine learning for hospitalized COVID-19 patients

**DOI:** 10.1101/2020.07.13.20150177

**Authors:** Miguel Marcos, Moncef Belhassen-García, Antonio Sánchez-Puente, Jesús Sampedro-Gomez, Raúl Azibeiro, P-Ignacio Dorado-Díaz, Edgar Marcano-Millán, Carolina García-Vidal, Maria-Teresa Moreiro-Barroso, Noelia Cubino-Bóveda, María-Luisa Pérez-García, Beatriz Rodríguez-Alonso, Daniel Encinas-Sánchez, Sonia Peña-Balbuena, Eduardo Sobejano-Fuertes, Sandra Inés, Cristina Carbonell, Miriam Lopez-Parra, Fernanda Andrade-Meira, Amparo López-Bernús, Catalina Lorenzo, Adela Carpio, David Polo-San-Ricardo, Miguel-Vicente Sánchez-Hernández, Rafael Borrás, Víctor Sagredo-Meneses, Pedro-L Sanchez, Alex Soriano, José-Ángel Martín-Oterino

## Abstract

**BACKGROUND:** Efficient and early triage of hospitalized Covid-19 patients to detect those with higher risk of severe disease is essential for appropriate case management.

**METHODS:** We trained, validated, and externally tested a machine-learning model to early identify patients who will die or require mechanical ventilation during hospitalization from clinical and laboratory features obtained at admission. A development cohort with 918 Covid-19 patients was used for training and internal validation, and 352 patients from another hospital were used for external testing. Performance of the model was evaluated by calculating the area under the receiver-operating-characteristic curve (AUC), sensitivity and specificity.

**RESULTS:** A total of 363 of 918 (39.5%) and 128 of 352 (36.4%) Covid-19 patients from the development and external testing cohort, respectively, required mechanical ventilation or died during hospitalization. In the development cohort, the model obtained an AUC of 0.85 (95% confidence interval [CI], 0.82 to 0.87) for predicting severity of disease progression. Variables ranked according to their contribution to the model were the peripheral blood oxygen saturation (SpO2)/fraction of inspired oxygen (FiO2) ratio, age, estimated glomerular filtration rate, procalcitonin, C-reactive protein, updated Charlson comorbidity index and lymphocytes. In the external testing cohort, the model performed an AUC of 0.83 (95% CI, 0.81 to 0.85). This model is deployed in an open source calculator, in which Covid-19 patients at admission are individually stratified as being at high or non-high risk for severe disease progression.

**CONCLUSIONS:** This machine-learning model, applied at hospital admission, predicts risk of severe disease progression in Covid-19 patients.

## Introduction

Since late 2019, a pneumonia outbreak caused by coronavirus SARS-CoV-2 began in the Chinese city of Wuhan and has evolved into a global pandemic.^1^ Clinical manifestations of patients with SARS-CoV-2 infection range from mild disease (e.g., only fever or cough) to critically ill cases with acute respiratory distress syndrome and septic shock. In a large report from the Chinese Center for Disease Control and Prevention, with 44415 cases, 36160 (81%) were described as mild, 6168 (14%) as severe, and 2087 (5%) as critical illness, with a mortality of 49% in the latter group.^2^ Due to this variability, several factors have been identified to predict increased severity, such as older age, neutrophilia, organ dysfunction, coagulopathy, or elevated D-dimer levels.^3^

Machine-learning is a subfield of computer science and statistics that has received growing interest in medicine, especially in infectious diseases, and has allowed to develop tools to predict clinical outcomes such as the occurrence of sepsis in intensive care units or the diagnosis of surgical site infection.^4^ Therefore, in this context of worldwide health emergency, early detection of patients who are likely to develop critical illness is of paramount importance and may aid in delivering proper care and optimizing use of limited intensive care resources.

For this purpose, we report here a machine-learning model able to predict risk of severity of disease progression in Covid-19 patients at the time of admission, developed and validated in two large cohorts of patients from two university hospitals, including easy-to-collect variables such as peripheral blood oxygen saturation (SpO2)/fraction of inspired oxygen (FiO2) ratio, age, estimated glomerular filtration rate, procalcitonin, C-reactive protein, updated Charlson comorbidity index and lymphocytes.

## METHODS

### Study Design and Data Sources

We conducted a training, validation an external-testing study on an intelligence-based machine-learning model,^5^ using clinical and laboratory features obtained at hospital admission. A data set from 918 confirmed Covid-19 patients from the University Hospital of Salamanca, Spain, was used for training and internal validation. For external testing we included 352 Covid-19 patients from another university hospital (Hospital Clinic of Barcelona, Spain).

Institutional approval was provided by the Ethics Committee of the University Hospital of Salamanca (2020/03/470) and the Comité Ètic d’Investigació Clínica of the Hospital Clínic of Barcelona (HCB/2020/0273), which waived the need for informed consent. All data set were anonymously analyzed, and the study was performed following current recommendation of the Declaration of Helsinki^6^.

### Task Definition

The aim of our study was to develop and validate a machine-learning model to predict, at the moment of hospital admission, the likelihood that a Covid-19 patient will die or require mechanical ventilation during hospitalization. A secondary objective was to deploy this model into a simple clinical digital application to facilitate its use in real time.

Input data (features) consists in demographic variables (including age and sex), individual comorbidities and Charlson Comorbidity Index, chronic medical treatment, clinical characteristics, physical examination parameters, and biochemical parameters available at hospital admission (Tables 1 and 2). As for the corresponding outcome (label), we defined severity of disease progression during hospitalization as the use of mechanical ventilation or death.

**Table 1.**
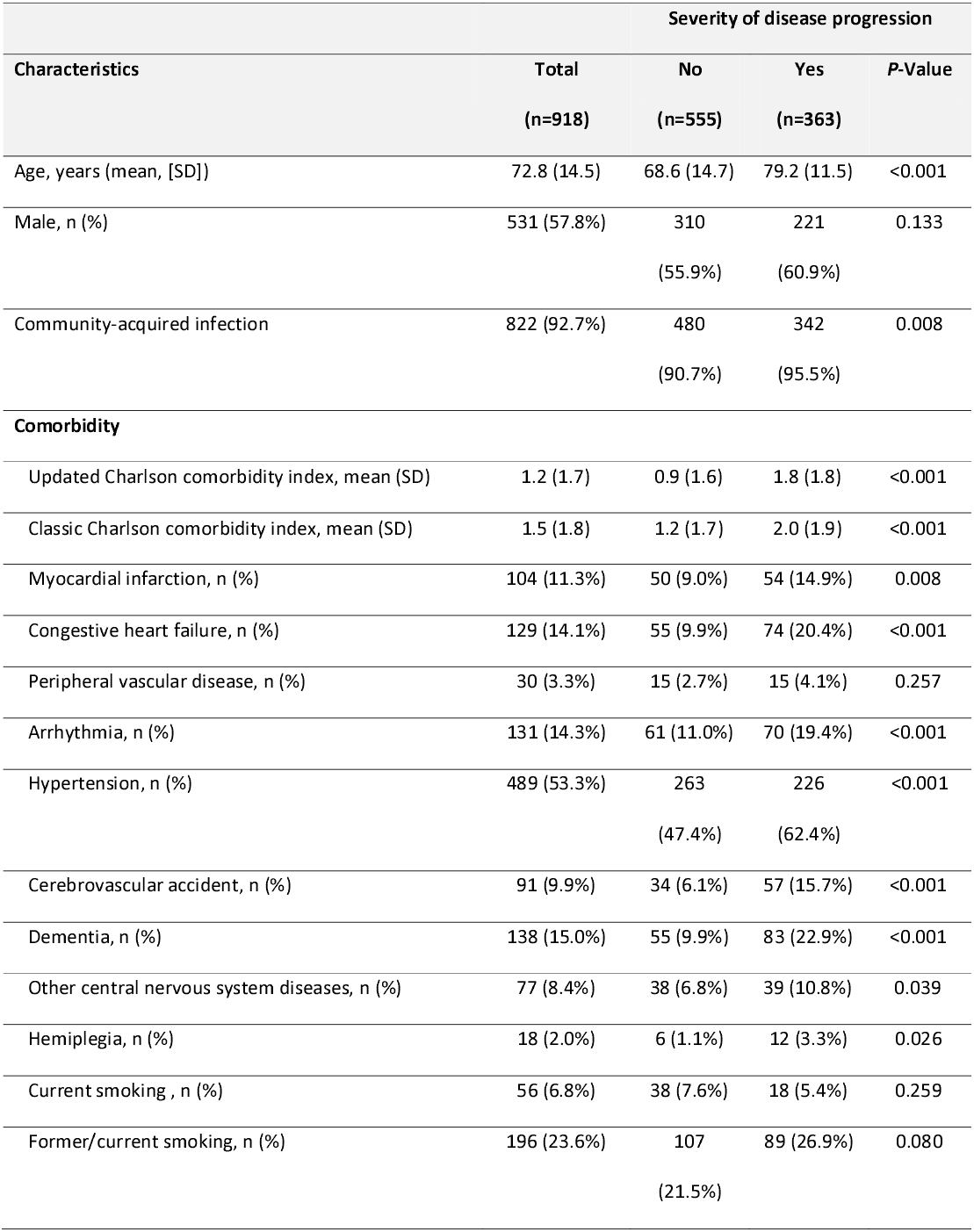
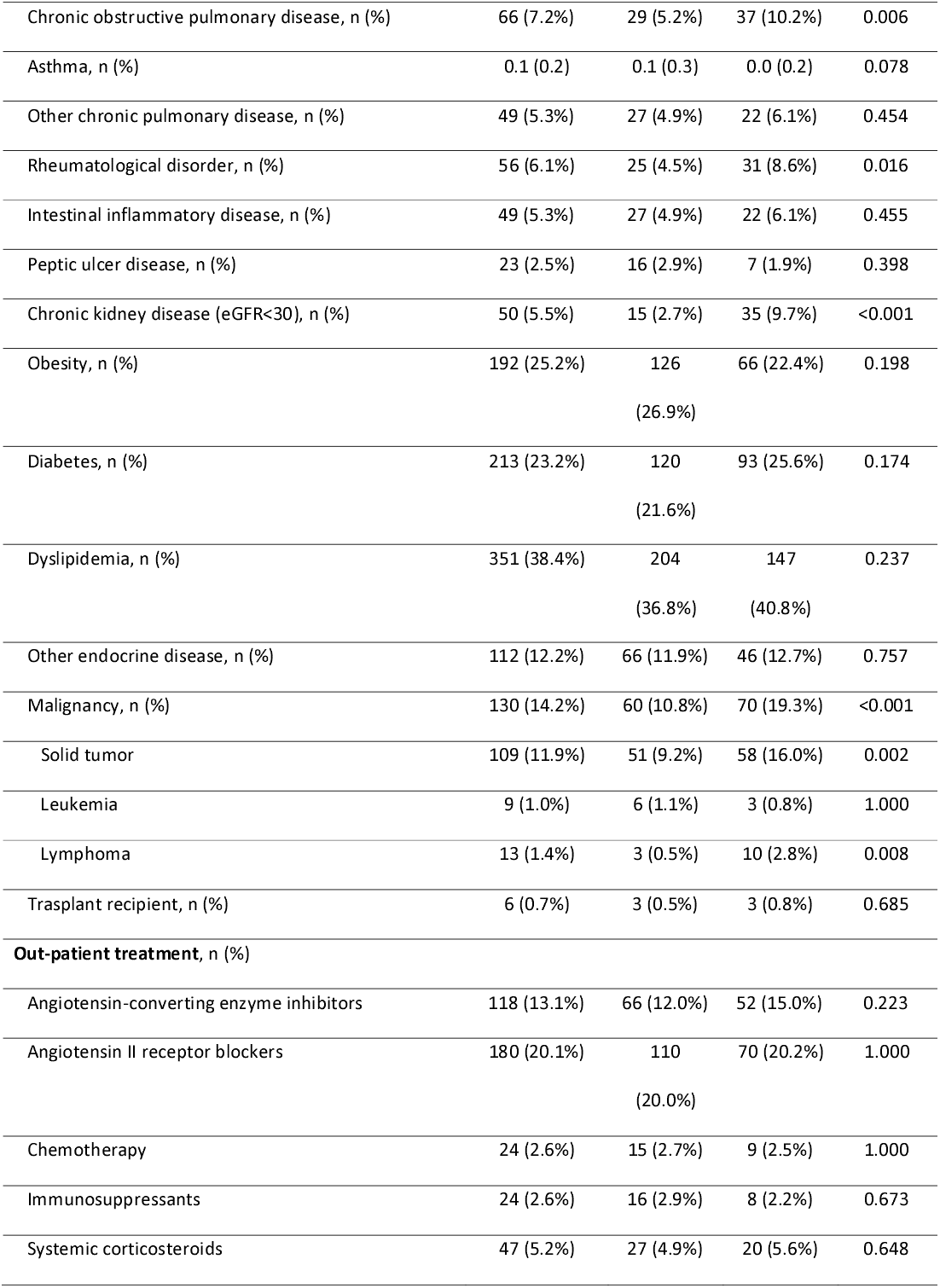
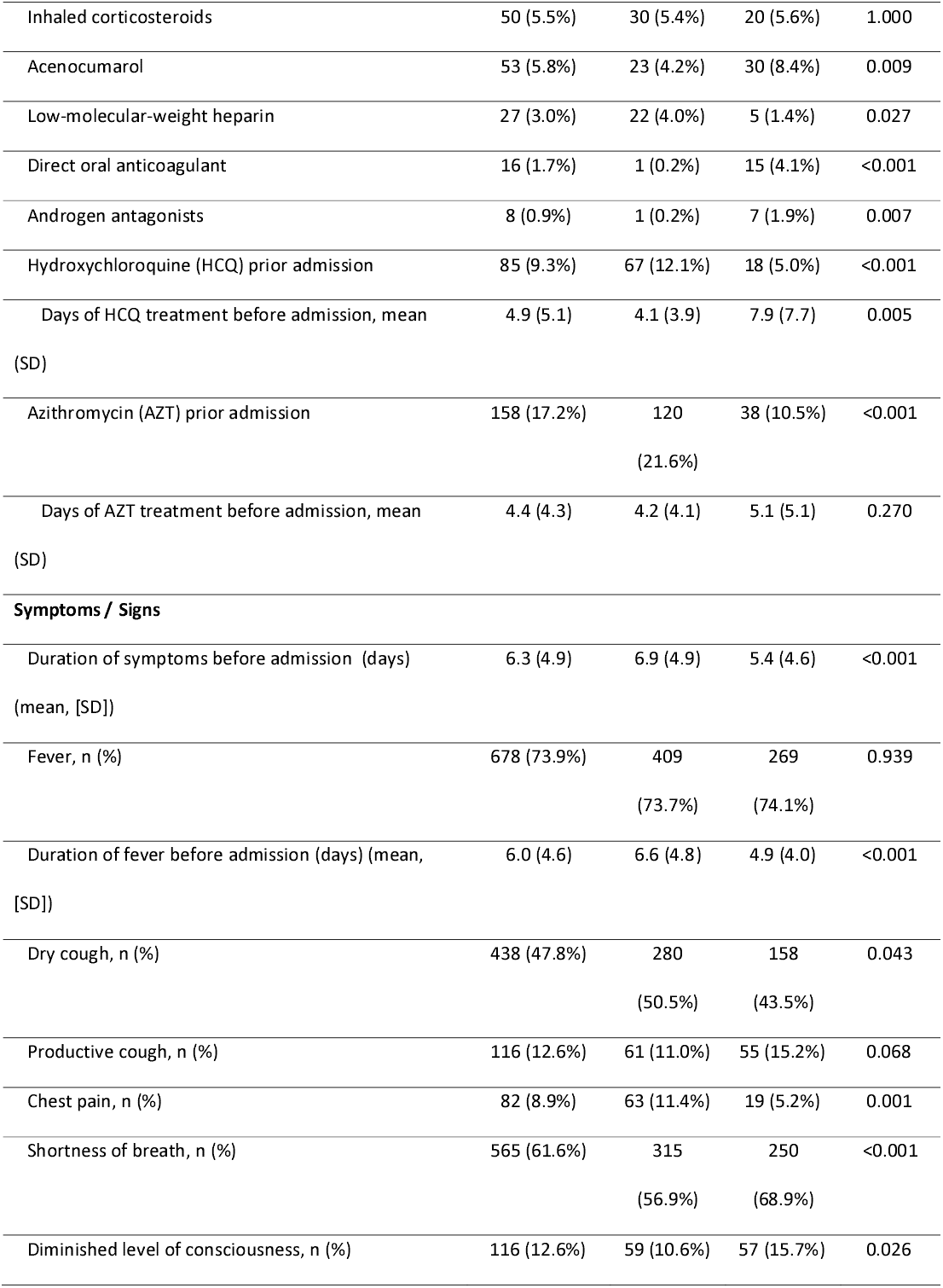
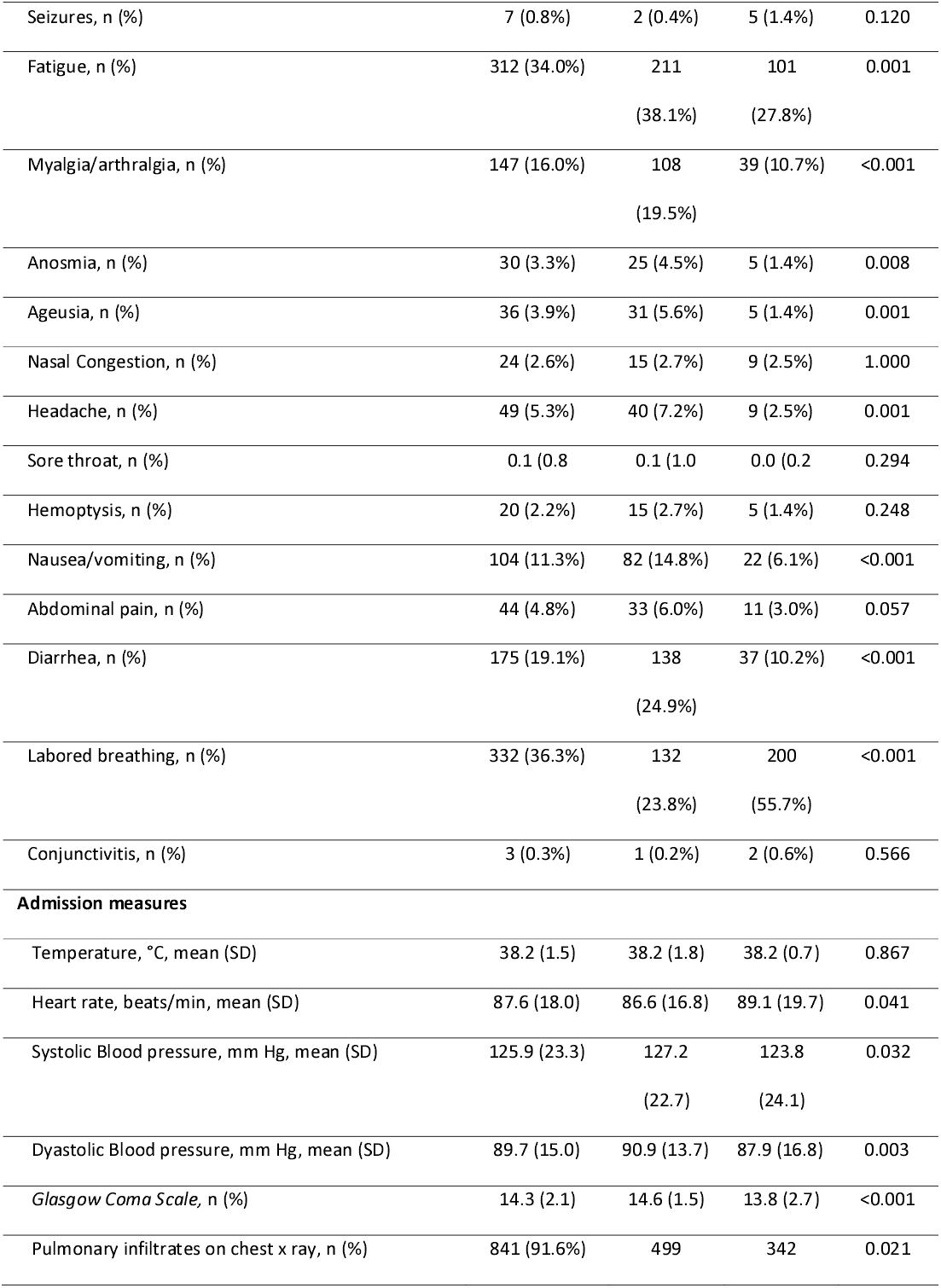
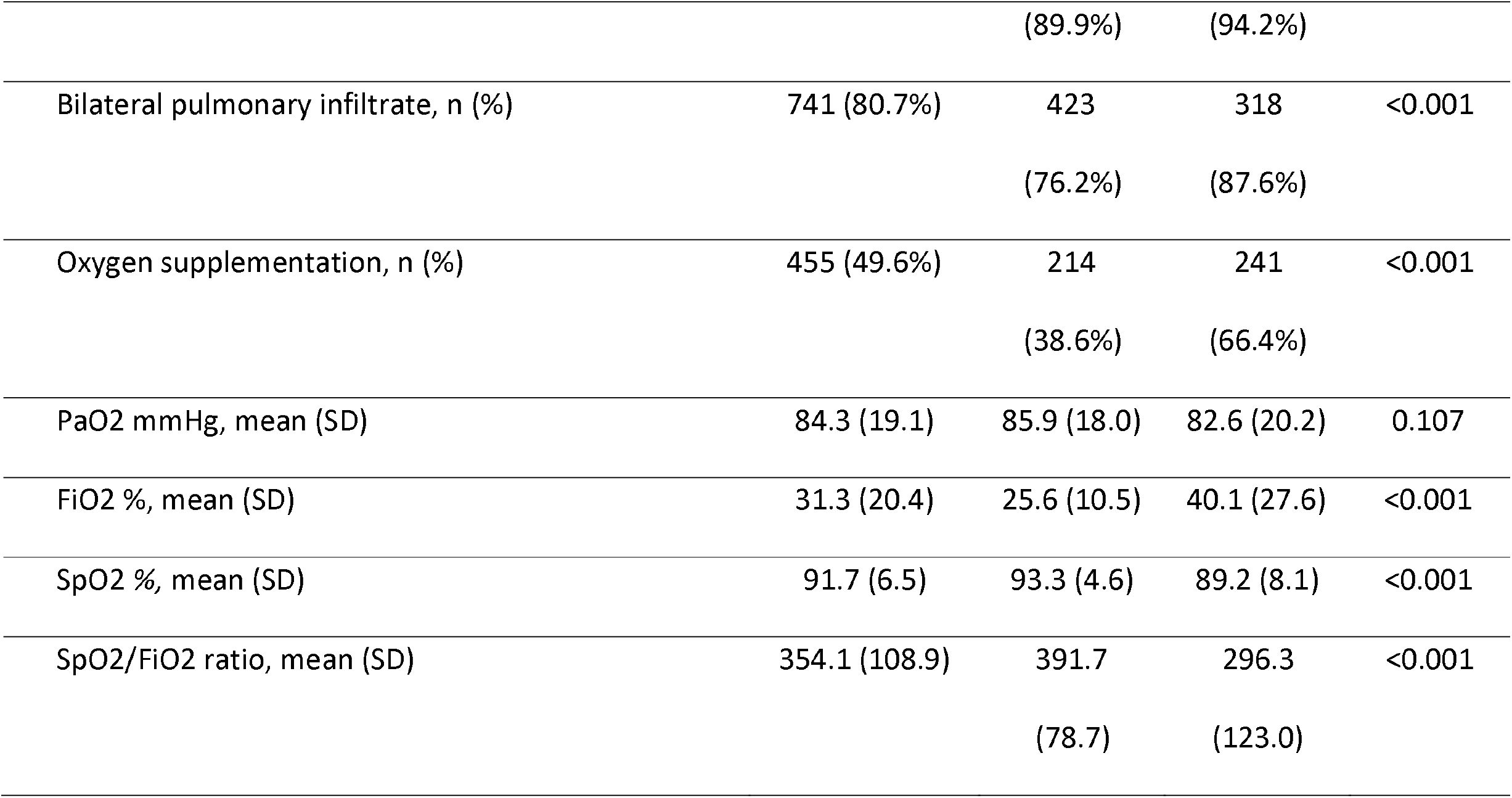
Admission characteristics of patients from internal validation cohort by outcome.

**Table 2.** Admission laboratory findings of patients from internal validation cohort by outcome.

| Characteristics, mean (SD) | Severity of disease progression |  |  |  |
| --- | --- | --- | --- | --- |
|  | Total | No | Yes | P-Value |
|  | (n=918) | (n=555) | (n=363) |  |
| Hemoglobin, g/dL | 13.6 (2.1) | 13.9 (2.0) | 13.2 (2.2) | <0.001 |
| Reticulocytes | 37.8 (20.8) | 34.5 (16.9) | 44.0 (25.7) | 0.031 |
| White blood cells count, x10<sup>9</sup> /L | 8.5 (14.1) | 8.4 (17.8) | 8.6 (4.2) | 0.826 |
| Neutrophil cell count, x10<sup>9</sup> /L | 6.2 (3.6) | 5.7 (3.2) | 7.1 (4.0) | <0.001 |
| Lymphocyte count, x10<sup>9</sup> /L | 1.7 (12.9) | 2.1 (16.5) | 1.0 (0.8) | 0.188 |
| Monocyte count, x10<sup>9</sup> /L | 0.5 (0.5) | 0.5 (0.6) | 0.5 (0.4) | 0.234 |
| Basophil count, x10<sup>9</sup> /L | 0.0 (0.0) | 0.0 (0.0) | 0.0 (0.0) | 0.010 |
| Platelet-to-lymphocyte ratio, % | 262.4 (226.6) | 245.0 (238.5) | 289.3 (204.1) | 0.004 |
| Neutrophil-to-lymphocyte Ratio, % | 8.2 (7.9) | 6.6 (5.7) | 10.6 (10.0) | <0.001 |
| Platelet count x10<sup>9</sup> /L | 209.0 (91.2) | 212.2 (92.1) | 204.0 (89.8) | 0.183 |
| Prothrombin time, % | 82.9 (22.0) | 85.9 (18.5) | 78.3 (25.8) | <0.001 |
| INR | 1.4 (2.0) | 1.3 (1.6) | 1.7 (2.4) | 0.010 |
| Activated partial thromboplastin time, s | 35.2 (8.3) | 35.1 (7.9) | 35.4 (8.8) | 0.686 |
| Fibrinogen levels, mg/dL | 637.3 (200.6) | 618.8 (193.9) | 666.2 (207.5) | 0.001 |
| D-dimer level, µg/mL | 2.9 (9.1) | 2.5 (9.2) | 3.4 (8.9) | 0.165 |
| ISTH-DIC score | 1.9 (1.2) | 1.6 (1.2) | 2.3 (1.2) | <0.001 |
| SOFA Score | 1.7 (1.9) | 1.1 (1.3) | 2.7 (2.1) | <0.001 |
| C-reactive protein, mg/dL | 13.2 (10.8) | 10.6 (9.0) | 17.2 (12.1) | <0.001 |
| Creatinine, mg/dL | 1.2 (0.7) | 1.1 (0.6) | 1.5 (0.9) | <0.001 |
| Bilirubin (total),mg/dL | 0.6 (0.5) | 0.6 (0.5) | 0.6 (0.4) | 0.439 |
| Ferritin, ng/mL | 1237.0 (1449.8) | 1052.9 (966.6) | 1610.9 (2069.6) | <0.001 |
| Glucose, mg/dL | 138.5 (63.8) | 130.1 (57.9) | 151.5 (70.2) | <0.001 |
| Urea, mg/dL | 58.7 (44.2) | 45.9 (33.6) | 78.4 (51.0) | <0.001 |
| Uric acid, mg/dL | 5.4 (2.5) | 4.8 (2.1) | 6.3 (2.7) | <0.001 |
| eGFR, mL/min/1.73 m<sup>2</sup> | 63.4 (25.6) | 71.7 (23.1) | 50.7 (23.9) | <0.001 |
| Calcium, mg/dL | 8.9 (0.7) | 9.0 (0.6) | 8.8 (0.7) | <0.001 |
| Magnesium, mmol/L | 2.1 (0.4) | 2.1 (0.3) | 2.2 (0.4) | 0.001 |
| Sodium, mmol/L | 137.8 (6.8) | 137.3 (5.6) | 138.7 (8.2) | 0.004 |
| Potassium, mmol/L | 4.1 (0.6) | 4.0 (0.5) | 4.2 (0.6) | <0.001 |
| Alanine Aminotransferase, U/L | 39.9 (72.5) | 38.4 (33.9) | 42.2 (108.3) | 0.451 |
| Aspartate Aminotransferase, U/L | 62.5 (175.7) | 49.3 (35.1) | 89.0 (299.0) | 0.012 |
| Alkaline phosphatase, U/L | 83.9 (68.4) | 80.8 (60.9) | 88.8 (78.6) | 0.092 |
| Gamma-glutamyltransferase, U/L | 72.2 (142.3) | 71.6 (108.8) | 73.2 (183.0) | 0.879 |
| Lactate dehydrogenase, U/L | 381.2 (175.1) | 345.3 (128.6) | 437.5 (218.5) | <0.001 |
| Proteins, g/L | 7.5 (0.6) | 7.5 (0.6) | 7.4 (0.7) | 0.001 |
| Albumin, g/L | 3.6 (0.4) | 3.8 (0.4) | 3.5 (0.4) | <0.001 |
| Creatine kinase, U/L | 225.9 (504.2) | 177.5 (364.4) | 304.0 (664.6) | 0.001 |
| Procalcitonin, ng/mL | 1.2 (6.1) | 0.7 (5.4) | 1.9 (6.9) | 0.018 |
| Bicarbonate, mEq/L | 25.2 (5.3) | 25.6 (4.7) | 24.7 (5.8) | 0.101 |
| Base excess, mEq/L | 1.0 (5.1) | 1.7 (4.3) | 0.2 (5.6) | 0.006 |

INR: International normalized ratio; International Society Thrombosis Hemostasis-Disseminated Intravascular Coagulation; SOFA, Sequential Organ Failure Assessment; eGFR, estimated glomerular filtration rate

### Data Preparation

The data was preprocessed by one-hot encoding multicategory features and completing missing values with the trimmed mean between 5-95 percentiles and mode of each continuous and categorical feature, respectively. The data was split in a train and validation data set following a 10-stratified fold cross-validation scheme with 10 repetitions^7^, in order to get a better assessment of the generalization performance of the classifiers.

### Training And Validation Of The Classification Machine-Learning Model

Three machine-learning classifiers typically used in data sets composed by heterogeneous features^8^ were trained: random forest,^9^ xgboost^10^ and regularized logistic regression. In the training phase, all models were optimized by fine tuning their hyperparameters with 10-fold cross-validation scheme and a grid search algorithm, configuring a nested cross-validation scheme to first perform this hyperparameter optimization and secondly internally evaluate the classifier. The fixed values of not optimized hyperparameters and the ranges of optimized ones for each classification and feature selection algorithm can be consulted in Table S1 in the Supplementary Data.

The code to develop the models was written in Python and open source libraries scikit-learn,^11^ xgboost and eli5 were used for the implementation of the machine-learning classifiers and cross-validation schemes. The code can be consulted at http://github.com/hus-ml/covid19salamanca-score

In order to better assess the clinical significance of our results, a real-world application of the model was evaluated with patients from a second tertiary university center, the Hospital Clinic of Barcelona.

### Evaluation Metrics

The differences in clinical, epidemiological and analytical variables between patients with and without severe disease progression at both hospitals were compared using χ2 tests for categorical variables and Student’s t-test for continuous variables.

The performance of the model was evaluated by calculating the area under the receiver-operating-characteristic curve (AUC) and its confidence interval for each prediction model^12,13^. The classification performance at particular cutoff thresholds based on the receiver-operating-characteristic curve were evaluated according to its sensitivity, specificity, positive predictive value, and negative predictive value.

### Severity Of Disease Classification Calculator

The developed machine-learning model was deployed in an open source calculator that can be run on a web application (https://covid19salamanca-score.herokuapp.com), in which Covid-19 patients at hospital admission can be individually stratified as high and non-high risk for severity of disease progression.

To develop a friendly calculator, the number of features used by the machine-learning model was reduced from 140 to less than 10. In order to ensure that all relevant clinical features were present a number of additional models were built, i.e. using combinations of death or death plus the use of mechanical ventilation as label and restricting the data set to older of 75 years of age, younger than 75 years of age or without age-restriction. The importance of each feature for these models was computed using Mean Decrease Accuracy.^9^ We tallied the number of times each feature appeared as one of the most important in a model and chose the most frequent features. Additionally, correlated features with similar importance were chosen by clinical significance and by their availability in the external data set. The new model developed with the selected features was validated to ensure similar results to the original one with all the features.

## Results

### Development cohort

Between March 1^st^ and April 23^rd^ 2020, among 918 patients that had been admitted at the University Hospital of Salamanca because of SARS-CoV-2 pneumonia, 363 patients (39.5%) died or required mechanical ventilation by May 15th (312 patients died and 82 required mechanical ventilation-31 of them finally died-) and 555 patients (60.5%) did not progress to critical illness and had been discharged by that date. Diagnosis was confirmed by RT-PCR assay from nasopharyngeal swab or immunochromatography assay in 859 and 59 patients, respectively. Table 1 shows features of patients of this cohort by severity of disease progression.

Concerning clinical variables, patients with severe disease progression were older (average age 79.2 years) and presented with a higher updated Charlson comorbidity index (mean value of 1.8). Overall, patients who developed critical illness had more cardiovascular and central nervous system diseases, and 35 out of 50 patients (70%) with chronic kidney disease had severe disease progression. Cancer was more prevalent in those with severity of disease progression (19.3% vs. 10.8%). Regarding clinical manifestations at admission, shortness of breath and labored breathing were present in 68.9% and 55.7% of the patients who progressed to severe disease, respectively. This group of patients had a significantly lower ratio of oxygen saturation as measured by pulse oximetry divided by the fraction of inspired oxygen (391.7 vs. 296.3) and 66.4% of them required oxygen supplement at admission whilst only 38.6% in the non-severe progression group.

Table 2 represents the laboratory findings at the time of admission by outcome. The patients with severity of disease progression presented at admission with neutrophilia, lymphopenia and higher levels of D-dimer, ferritin, C-reactive protein, procalcitonin and fibrinogen. The critically ill group patients had altered renal function at admission, measured by increased urea and creatinine levels and reduced estimated glomerular filtration rate.

### Risk Model Performance

In order to develop the risk model, we first selected all variables included in the tables 1 and 2. Using all the cohort patients and variables, the best model obtained in the internal cross-validation an AUC of 0.86 (CI: 0.83-0.88). With the aim of developing a more user-friendly application and according to the described methodology, we identified 7 variables present in all models with independent prognostic significance: peripheral blood oxygen saturation (SpO2)/fraction of inspired oxygen (FiO2) ratio, age, estimated glomerular filtration rate, procalcitonin, C-reactive protein, updated Charlson comorbidity index and lymphocytes. By restricting to these 7 variables, out of the 3 trained machine-learning classifiers, the best classifier achieved a highest mean AUC of 0.85 (CI: 0.82-0.87) from our development cohort without significant difference among them (Figure 1).

**Figure 1.**
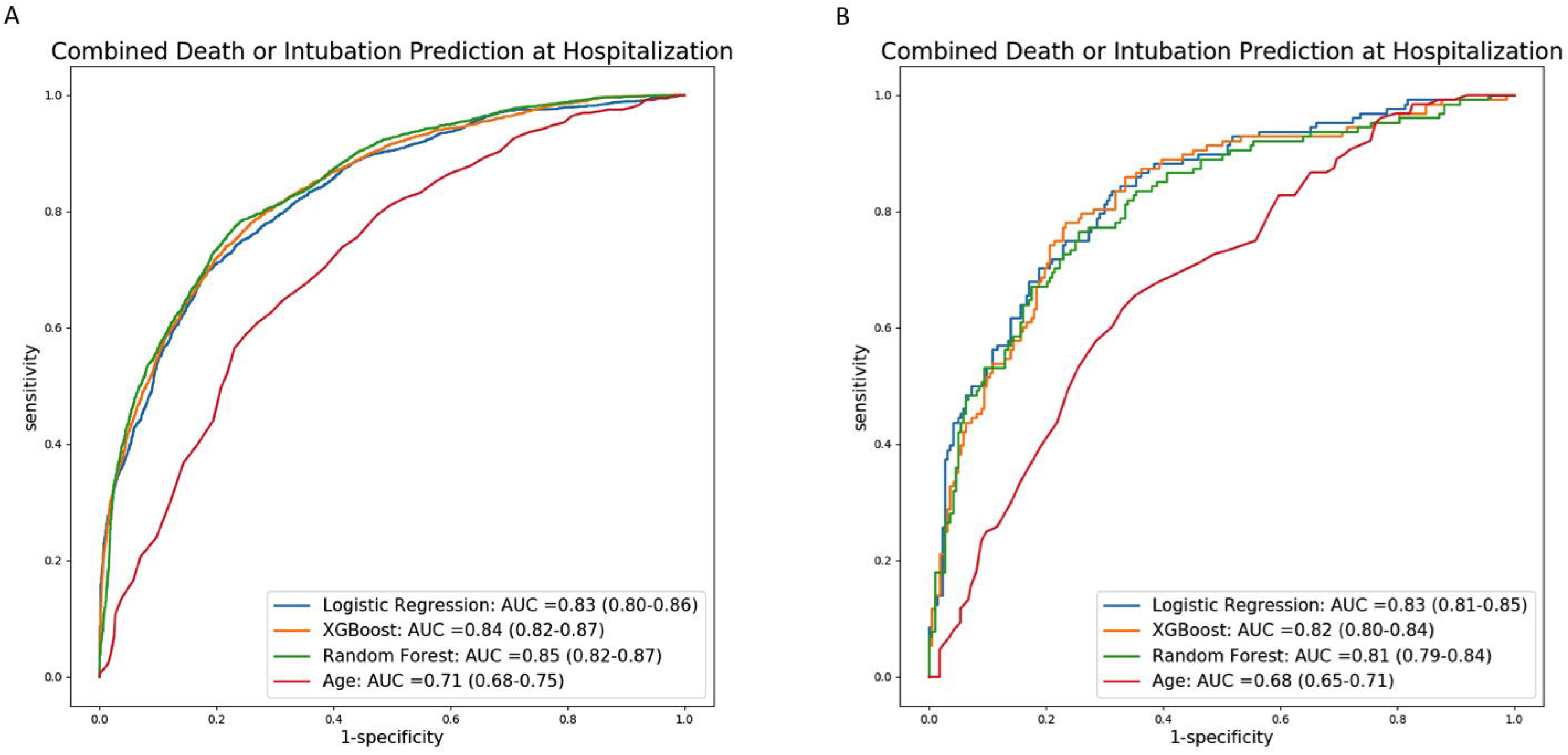
Receiver operating characteristic curves of the machine-learning model for the different classification algorithms. Panel A shows the internal crossvalidation results and panel B shows the external testing results. The results of a risk classification only based on age are also shown for comparison

### External Testing cohort

Between February 15^th^ and April 28^th^ 2020, 352 patients were admitted at the Clinic Hospital of Barcelona because of their first episode of SARS-CoV-2 pneumonia confirmed by RT-PCR assay from nasopharyngeal swab. Among them, 128 (36.3%) patients developed critical illness (64 died and 77 required mechanical ventilation-13 of them finally died-) and 224 (63.6%) did not and were discharged by May 20th. The baseline characteristics and laboratory findings at the time of admission in this external testing cohort are represented in the Table 3. Patients with severity of disease progression were older (median age of 68.7), with lower SpO2/FiO2%, lower glomerular filtration rate and higher procalcitonin and C-reactive protein values. In addition, they presented with lower lymphopenia count and higher updated CCI scores.

**Table 3.** Admission demographic and clinical characteristics of patients from external testing cohort by outcome.

| Characteristics | Severity of disease progression |  |  |  |
| --- | --- | --- | --- | --- |
|  | Total | No | Yes | *P*-value |
|  | (n=352) | (n=224) | (n=128) |  |
| Male, n (%) | 235 (66.8%) | 148 (66.1%) | 87 (68.0%) | 0.726 |
| Age, mean (SD) | 62.9 (14.6) | 59.5 (15.1) | 68.7 (11.6) | <0.001 |
| SpO<sub>2</sub>/FiO<sub>2</sub>, %/%, mean (SD) | 3.80 (1.09) | 4.18 (0.69) | 3.11 (1.32) | <0.001 |
| eGFR, mL/min/1.73 m<sup>2</sup>, mean (SD) | 77.6 (24.4) | 83.0 (21.6) | 67.9 (26.3) | <0.001 |
| Procalcitonin, ng/mL, mean (SD) | 0.6 (2.7) | 0.3 (0.8) | 1.1 (4.3) | 0.035 |
| Lymphocyte count, x10<sup>9</sup> /L, mean (SD) | 0.8 (0.4) | 0.9 (0.4) | 0.7 (0.4) | <0.001 |
| C-reactive protein, mg/L, mean (SD) | 11.9 (14.7) | 8.6 (6.9) | 17.9 (21.7) | <0.001 |
| Updated CCI, mean (SD) | 0.8 (1.0) | 0.6 (0.9) | 1.1 (1.1) | <0.001 |

FiO<sub>2</sub>, fraction of inspired oxygen; SpO<sub>2</sub>, arterial oxygen saturation measured by pulse oximetry; eGFR, estimated glomerular filtration rate; CCI: Charlson Comorbidity Index

The three trained classifiers restricted to the 7 most relevant variables were externally validated on this cohort. In this case, the best classifier obtained a mean AUC of 0.83 (CI: 0.81-0.85), again without significant differences respect to the other classifiers (Figure 1) and very consistent with the results obtained in the development cohort.

The relative contribution to the AUC of each feature both in the development and testing populations are shown in Table 4. In both cohorts, SpO2/FiO2 and C-reactive protein were the best predictors of critical evolution of disease, while procalcitonin and lymphocyte count showed lower contribution to the prediction.

**Table 4.** Relative importance of each variable according to MDA, scaled to the most important one.

|  | Logistic Regression | Random Forest | XGBoost |
| --- | --- | --- | --- |
| SpO2/FiO2 | 1 | 1 | 1 |
| C-reactive protein | 0.353 | 0.218 | 0.102 |
| eGFR | 0.381 | 0.328 | 0.200 |
| Age | 0.26 | 0.291 | 0.150 |
| Updated CCI | 0.19 | 0.185 | 0.076 |
| Lymphocyte count | 0.039 | 0.183 | 0.097 |
| Procalcitonin | 0.003 | 0.213 | 0.179 |

MDA, Mean Decrease Accuracy; SpO2, arterial oxygen saturation measured by pulse oximetry; FiO2, fraction of inspired oxygen; eGFR, estimated glomerular filtration rate; CCI: Charlson comorbidity index

### Calculator application

The 7-variable model based on the regularized logistic regression, which obtained the best result in the external testing cohort, has been deployed in an open-source web calculator (https://covid19salamanca-score.herokuapp.com/) to predict the risk of severity of disease progression, with the possibility of selecting different cut-off thresholds to individualize the definition of high risk of severity of disease progression depending on the hospital resources. As example, a high availability resource cut-off threshold is estimated to obtain in the internal validation cohort a sensitivity of 0.90 and specificity of 0.52 for detecting high-risk patients, which result in the identification of 2 groups of patients representing the 64.6% and 35.4% of the cohort with 55.1% and 11.2% of them developing severe disease, respectively (Figure S1). These values of sensitivity and specificity are susceptible to change in populations with different risk distributions (e.g., younger populations) or if there are other pre-admission criteria that skew the population. As a consequence, this high availability resource cut-off threshold, evaluated on the external setting cohort (younger population), identified groups including the 39.2% and 60.8% of the population with 65.9% and 17.3% of them developing severe disease, respectively.

## Discussion

In this study, we have developed and validated, through machine-learning, a clinical risk score to predict at the moment of hospital admission by Covid-19 pneumonia, the risk of mechanical ventilation or death. This score is also provided as an open-source web-based calculator, which allows clinicians to estimate an individual Covid-19 patient risk and make decisions based on availability of resources for critical patients and patient overload.

This score includes several common and readily available variables that may be collected at admission in most hospitals. Both development and testing cohorts of patients are representative series for gaining insights into the prediction of disease severity in Covid-19 patients because both are university institutions, patients were in charge of Infectious Diseases/Internal Medicine Departments and treatment protocols were quite homogeneous due to the recommendations of the Spanish Agency of Medicines and Medical Devices (AEMPS). Further, the selected time frame corresponds to the peak Covid-19 incidence and excess mortality in Spain.

As far as the variables included in the risk model here presented, age has been described as one of the main risk factors predicting severity and inpatient mortality in Covid-19 and other scores have also included this variable^2,14^. Concerning comorbidities, although the exact type and number of comorbidities posing more risk for adverse outcomes is still unknown^2,14^, our analysis has shown that updated Charlson comorbidity index was the most powerful variable to integrate and combine comorbidities at admission and resulted better than individual variables, such as hypertension or heart failure, or the classical Charlson index. Considering that the updated Charlson index is an improved and more parsimonious prognostic score than the classical one and has been previously described as a prognostic tool in many settings, including infectious diseases,^15^ this score may therefore serve to adjust for comorbidity in other Covid-19 studies.

The ratio of oxygen saturation as measured by pulse oximetry divided by the fraction of inspired oxygen is a simple measure, which has been previously used in the setting of acute respiratory distress syndrome instead of more complex variables,^16^ and thus can be evaluated in each patient with Covid-19 pneumonia to help identify patients at higher risk of severe disease.

Regarding laboratory variables, decreased estimated glomerular filtration rate and increased acute phase reactants like procalcitonin or C-reactive protein are associated with higher risk of severe disease. Although renal disease is part of the Charlson index as a comorbid disease, decreased estimated glomerular filtration rate may indicate not only the presence of this comorbidity but also acute kidney injury due to disease severity (e.g., septic shock). Therefore, it is a simple variable to assess severity of disease progression at the time of first visit. Indeed, kidney disease as a predictor of increased Covid-19 inpatient mortality rate has been previously described in a single-center study in China.^17^

Increased levels of C-reactive protein and its association with prognosis and severity in Covid-19 have been reported and correlated with pro-inflammatory response.^18^ Disease severity has also been linked with increased procalcitonin levels and described in some series although its elevation might be likely associated with the presence of bacterial superinfection.^19^ Low lymphocyte count has already been linked to poorer outcomes in Covid-19 inpatients and other viral infections such as influenza^18,20^. In addition, lymphopenia may play a pathogenic role in this disease due to a decrease of specific lymphocyte subpopulations and tissue infiltration^21^.

The score presented here exhibits a very good performance and accuracy, as well as excellent validation in the testing cohort with an easy-to-use web interface. Most previous scores have been developed and validated among Chinese Covid-19 patients^14,18^ and this is, in our knowledge, the first risk model developed with machine-learning methodology in Caucasian population including clinical and analytical variables at admission and therefore, it can be more applicable in Western countries. We would also like to highlight the possibility of the open-source web calculator to select different sensitivity cut-off thresholds to classify patients depending on health-care resources and population risk distributions. This possibility will clearly improve the efficiency of triage of Covid-19 patients at hospital admission through a real-time, automated and personalized method that would also take into account hospital intensive care unit availability within this pandemic situation.

This study, to assure uniformity, focused solely on patients admitted in a university hospital after an emergency department visit and did not include those Covid-19 cases managed in the outpatient setting. However, it would be optimal to validate this risk score at the time of first evaluation by family physicians to potentially identify patients at risk of progressive disease and thus allow early hospital derivation.

In summary, this risk model represents a reliable system that uses widely available clinical and laboratory parameters at hospital admission. The application of machine-learning methods has led to better prediction of the outcome for the identification of Covid-19 inpatients that will likely develop progressive disease after admission.

## Data Availability

Code to develop machine learning model is available.

http://github.com/hus-ml/covid19salamanca-score

https://covid19salamanca-score.herokuapp.com

## Acknowledgments

We acknowledge to Maria-Victoria Mateos and Jose-Ramon González-Porras from the Hematology Department of the University Hospital of Salamanca for her contribution with the analysis and writing of this manuscript as well as to all doctors and healthcare personnel involved in the COVID-19 Working Team from the Internal Medicine Department and others departments of the University Hospital of Salamanca: José-Ignacio Martín-González, José-Ignacio Madruga-Martín, María-José García-Rodríguez, Ángela Romero-Alegría, Nora Gutiérrez-San Pedro, Leticia Moralejo-Alonso, José-Ignacio Herrero-Herrero, Antonio Chamorro, Mercedes Martín-Ordiales, Celestino Martín-Álvarez, Guillermo Hernández, María Sánchez-Ledesma, María-José Sánchez-Crespo, Felipe Álvarez-Navia, Patricia Araoz-Sánchez, Judit Aparicio-García, Jacinto Herráez, Ronald Macías, Alejandro Rolo, Juan-Francisco Soto, Laura Manzanedo, Luis Seisdedos, Juan-Miguel Manrique, Alfredo-Javier Collado, Sandra Rodríguez, Ana Rodríguez, Silvia Ojea, Laura Burgos, Carlos Reina, Eugenia López, José-María Bastida, María Díez-Campelo, Alberto Hernández-Sánchez, Luis-Mario Vaquero, Ignacio Martín, Cristina de-Ramón, Estefanía Pérez, Borja Puertas, Daniel Presa, Ana Yeguas, Ana África, María-Victoria Coral, Rosa Tejera, Laura Gil, Javier Fernández, Elisa Acosta, Fátima Bouhmir, Sonia Pastor, Marta Fonseca, María-de-los-Ángeles Pérez-Nieto, Ernesto Parras, María Cartagena, Víctor Barreales, Óscar Humberto, María Bartol, Olga Compán, Ana Ramón, Raquel Rodríguez, Silvia Ruiz, Sonsoles Garrosa, Alexis-Alan Rodrigo, Sara Alonso, Raquel Domínguez, Felipe Peña, María García-Duque, Ángela Romero, Ana Menéndez, and all the staff members. We want also to thank all COVID-19 patients admitted at the University Hospital of Salamanca and their families.

We acknowledge to the Infectious Diseases’ Research Group of the Hospital Clínic of Barcelona: Albiach L, Agüero D, Ambrosioni J, Bodro M, Blanco JL, Cardozo C, Chumbita M, De la Mora L, García-Alcaide F, García-Pouton N, González-Cordón A, Hernández-Meneses M, Inciarte A, Laguno M, Leal L, Linares L, Macaya I, Mallolas J, Martínez E, Martínez M, Meira F, Miró JM, Mensa J, Moreno A, Moreno A, Moreno-García E, Morata L, Martínez JA, Puerta-Alcalde P, Rico V, Rojas J, Solá M, Tomé A, Torres B, Torres M, and all the staff members. We also acknowledge to the Medical Intensive Care Unit: Adrian Téllez, Sara Fernández, Pedro Castro, Josep M Nicolás, and all the staff members.

## Data sharing statement

Relevant anonymised patient level data are available on reasonable request. Code to develop machine learning model is already available.

## REFERENCES

1. Lu RJ, Zhao X, Li J, et al. Genomic characterisation and epidemiology of 2019 novel coronavirus: implications for virus origins and receptor binding. Lancet 2020;395:565–74.

2. Wu Z,McGoogan JM. Characteristics of and important lessons from the coronavirus disease 2019 (COVID-19) outbreak in China: summary of a report of 72 314 cases from the Chinese Center for Disease Control and Prevention. JAMA. 2020;24.

3. Yang XB, Yu Y, Xu JQ, et al. Clinical course and outcomes of critically ill patients with SARS-CoV-2 pneumonia in Wuhan, China: a single-centered, retrospective, observational study. Lancet Respiratory Medicine 2020;8:475–81.

4. Peiffer-Smadja N, Rawson TM, Ahmad R, et al. Machine learning for clinical decision support in infectious diseases: a narrative review of current applications. Clinical Microbiology and Infection 2020;26:584–95.

5. Rajkomar A, Dean J, Kohane I. Machine Learning in Medicine. N Engl J Med 2019;380:1347–58.

6. World Med A. World Medical Association Declaration of Helsinki Ethical Principles for Medical Research Involving Human Subjects. Jama-Journal of the American Medical Association 2013;310:2191–4.

7. Rodriguez JD, Perez A, Lozano JA. Sensitivity analysis of kappa-fold cross validation in prediction error estimation. IEEE Trans Pattern Anal Mach Intell 2010;32:569–75.

8. Dorado-Diaz PI, Sampedro-Gomez J, Vicente-Palacios V, Sanchez PL. Applications of Artificial Intelligence in Cardiology. The Future is Already Here. Revista Espanola De Cardiologia 2019;72:1065–75.

9. Breiman L. Random forests. Machine Learning 2001;45:5–32.

10. Chen TQ, Guestrin C, Assoc Comp M. XGBoost: A Scalable Tree Boosting System. Kdd’16: Proceedings of the 22nd Acm Sigkdd International Conference on Knowledge Discovery and Data Mining 2016:785-94.

11. Pedregosa F, Varoquaux G, Gramfort A, et al. Scikit-learn: Machine Learning in Python. Journal of Machine Learning Research 2011;12:2825–30.

12. Bouckaert R. Choosing Between Two Learning Algorithms Based on Calibrated Tests 2003.

13. Nadeau C, Bengio Y. Inference for the generalization error. Machine Learning 2003;52:239–81.

14. Liang W, Liang H, Ou L, et al. Development and Validation of a Clinical Risk Score to Predict the Occurrence of Critical Illness in Hospitalized Patients With COVID-19. JAMA Internal Medicine 2020.

15. Ternavasio-de la Vega HG, Castano-Romero F, Ragozzino S, et al. The updated Charlson comorbidity index is a useful predictor of mortality in patients with Staphylococcus aureus bacteraemia. Epidemiology and Infection 2018;146:2122–30.

16. Schmidt MFS, Gernand J, Kakarala R. The use of the pulse oximetric saturation to fraction of inspired oxygen ratio in an automated acute respiratory distress syndrome screening tool. Journal of Critical Care 2015;30:486–90.

17. Cheng Y, Luo R, Wang K, et al. Kidney disease is associated with in-hospital death of patients with COVID-Covid-19. Kidney Int. 2020;97:829–838

18. Yan L, Zhang H-T, Goncalves J, et al. An interpretable mortality prediction model for COVID-19 patients. Nature Machine Intelligence 2020;2:283–8.

19. Lippi G, Plebani M. Procalcitonin in patients with severe coronavirus disease 2019 (COVID-19): A meta-analysis. Clinica Chimica Acta 2020;505:190–1.

20. Lalueza A, Folgueira D, Diaz-Pedroche C, et al. Severe lymphopenia in hospitalized patients with influenza virus infection as a marker of a poor outcome. Infectious Diseases 2019;51:543–6.

21. Chen G, Wu D, Guo W, et al. Clinical and immunological features of severe and moderate coronavirus disease 2019. The Journal of clinical investigation 2020;130:2620–9.

